# CMV seropositivity is a potential novel risk factor for severe COVID-19 in non-geriatric patients

**DOI:** 10.1101/2021.12.22.21268268

**Authors:** Simone Weber, Victoria Kehl, Johanna Erber, Karolin I. Wagner, Ana-Marija Jetzlsperger, Theresa Burrell, Kilian Schober, Philipp Schommers, Max Augustin, Claudia S. Crowell, Markus Gerhard, Christof Winter, Christoph D. Spinner, Ulrike Protzer, Dieter Hoffmann, Elvira D’Ippolito, Dirk H Busch

**Author notes:** co-first authors (Simone Weber, Victoria Kehl). co-senior authors (Elvira D’Ippolito, Dirk H Busch). **CORRESPONDING AUTHOR** Prof. Dr. med. Dirk H. Busch, Institute for Medical Microbiology, Immunology and Hygiene, Technical University of Munich, Trogerstr. 30, 81675 Munich, Germany.

## Abstract

**Background:** COVID-19 has so far affected more than 250 million individuals worldwide, causing more than 5 million deaths. Several risk factors for severe disease have been identified, most of which coincide with advanced age. In younger individuals, severe COVID-19 often occurs in the absence of obvious comorbidities. Guided by the finding of cytomegalovirus (CMV)-specific T cells with some cross-reactivity to SARS-CoV-2 in a COVID-19 intensive care unit (ICU) patient, we decided to investigate whether CMV seropositivity is associated with severe or critical COVID-19.

**Methods:** National German COVID-19 bio-sample and data banks were used to retrospectively analyze the CMV serostatus of patients who experienced mild (n=101), moderate (n=130) or severe to critical (n=80) disease by CMV IgG serology. We then investigated the relationship between disease severity and CMV serostatus via statistical models.

**Results:** Non-geriatric patients (< 70 years) with severe COVID-19 were found to have a very high prevalence of CMV-seropositivity, while CMV status distribution in individuals with mild disease was similar to the prevalence in the German population; interestingly, this was not detectable in older patients. Prediction models support the hypothesis that the CMV serostatus might be a strong biomarker in identifying younger individuals with a higher risk of developing severe COVID-19.

**Conclusions:** We identified ‘CMV-seropositivity’ as a potential novel risk factor for severe COVID-19 in non-geriatric individuals in the studied cohorts. More mechanistic analyses as well as confirmation of similar findings in cohorts representing the currently most relevant SARS-CoV-2 variants should be performed shortly.

## INTRODUCTION

Despite world-wide vaccination efforts, another wave of severe acute respiratory syndrome coronavirus 2 (SARS-CoV-2) infections is currently rapidly emerging in many countries in the northern hemisphere, bringing hospital capacities to their limits.

In the meantime, it has been well documented that individuals of advanced age and/or with certain risk factors, such as cardiovascular or pulmonary diseases, obesity as well as male sex, have a higher mortality rate in the context of SARS-CoV-2 infection^1–3^. Although multiple risk factors for severe COVID-19 disease have been identified, there seems to be a broad spectrum of disease penetrance; in addition, younger individuals with severe disease sometimes do not show any of the known risk factors. As such, the reasons for the development of severe symptoms and subsequent need for intensive care unit (ICU) admission in many patients remain unclear.

In a prior study, we investigated the phenotype of SARS-CoV-2-specific T cells in severe COVID-19 patients who required invasive mechanical ventilation, and identified T cell receptors (TCRs) specifically recognizing and reacting to the spike protein of the virus^4^. Re-expression of the identified TCRs in primary human T cells^5^ allowed us to characterize the antigen reactivity profile of these SARS-CoV-2 reactive T cells in more detail. To our surprise, in follow up experiments we could identify a strong and robust cytokine response to human Cytomegalovirus (CMV) pp65 peptide mix in different TCRs specific to SARS-CoV-2 S-protein derived from an ICU COVID-19 patient (Supplementary Fig.1 a-b).

CMV is a herpesvirus that causes latently persisting infection and is transmitted through body fluids such as breastmilk or saliva. The prevalence varies geographically and is also associated with socioeconomic status^6,7^ – the prevalence in Low-to-Middle-Income-Countries is generally higher than in High-Income countries. CMV seropositivity is furthermore associated with cardiovascular comorbidities as well as a higher incidence of thromboembolic events^8–11^, which have already been linked to an increased risk for severe COVID-19 or have been shown to be a complication of SARS-CoV-2 infection^12^. While primary and latent CMV infections in immunocompetent individuals do not cause major symptoms, CMV (re-)activation is a feared complication in immunocompromised patients and new-borns^13–15^. Recently, a few cases of CMV reactivation in the setting of severe COVID-19 have been reported^16–19^.

CMV infection is also known to reshape the immune repertoire by creating an inflationary memory T cell response that can occupy a large fraction of the overall T cell pool^20,21^, creating so-called ‘memory inflation’^22^. This phenomenon becomes more prominent with increasing age, and CMV seropositivity has been linked to impaired immune responses to other infections as well as to vaccination by immunosenescence, especially in older individuals^15,23–25^. Therefore, it was speculated that the development of effective T cell responses upon infection with SARS-CoV-2 could be strongly dampened by CMV-driven immunosenescence^26,27^, which might at least in part explain the high prevalence of severe disease in the elderly (>80 years).

Overall, the identification of SARS-CoV-2/CMV cross-reactive T cells, the known impact of CMV infection on the immune system, as well as the first reports on CMV reactivation during severe COVID-19 guided us to investigate whether CMV seropositivity is associated with severe COVID-19.

To address this question, CMV-serostatus was retrospectively analysed via the measurement of CMV IgG titers in cohorts of patients with mild to severe COVID-19 disease. To our surprise, these data show that CMV seropositivity is strongly associated with development of severe disease in individuals younger than 70 years. We could not identify such a pattern in elderly individuals (> 70 years).

## RESULTS

To investigate the possible influence of an individual’s CMV status on the course of COVID-19, we analyzed serum samples from SARS-CoV-2 infected individuals who experienced different disease severity. CMV IgG was measured on a total of 311 individuals with either mild (not admitted to the hospital, n=101, median age 50-59), moderate (hospitalized but no ICU admission, n=130, median age 60-69) or severe to critical (ICU, n=80, median age 70-79) disease. Where available, data on pre-existing comorbidities were also collected (Table 1). As expected, patients who experienced more severe symptoms were of older age and/or more likely to suffer from comorbidities, with almost 90% of ICU patients being affected by at least one comorbidity (Table 1). In line with this observation as well as with existing evidence, we also found age and comorbidities to be strong risk factors for severe COVID-19 (Table 2, univariate analyses). Furthermore, prevalence of these known comorbidities clearly rose with increasing age in our cohort (Supplementary Fig. 2A), thus supporting the relationship of these two variables in predicting COVID-19 outcome.

**Table 1:**
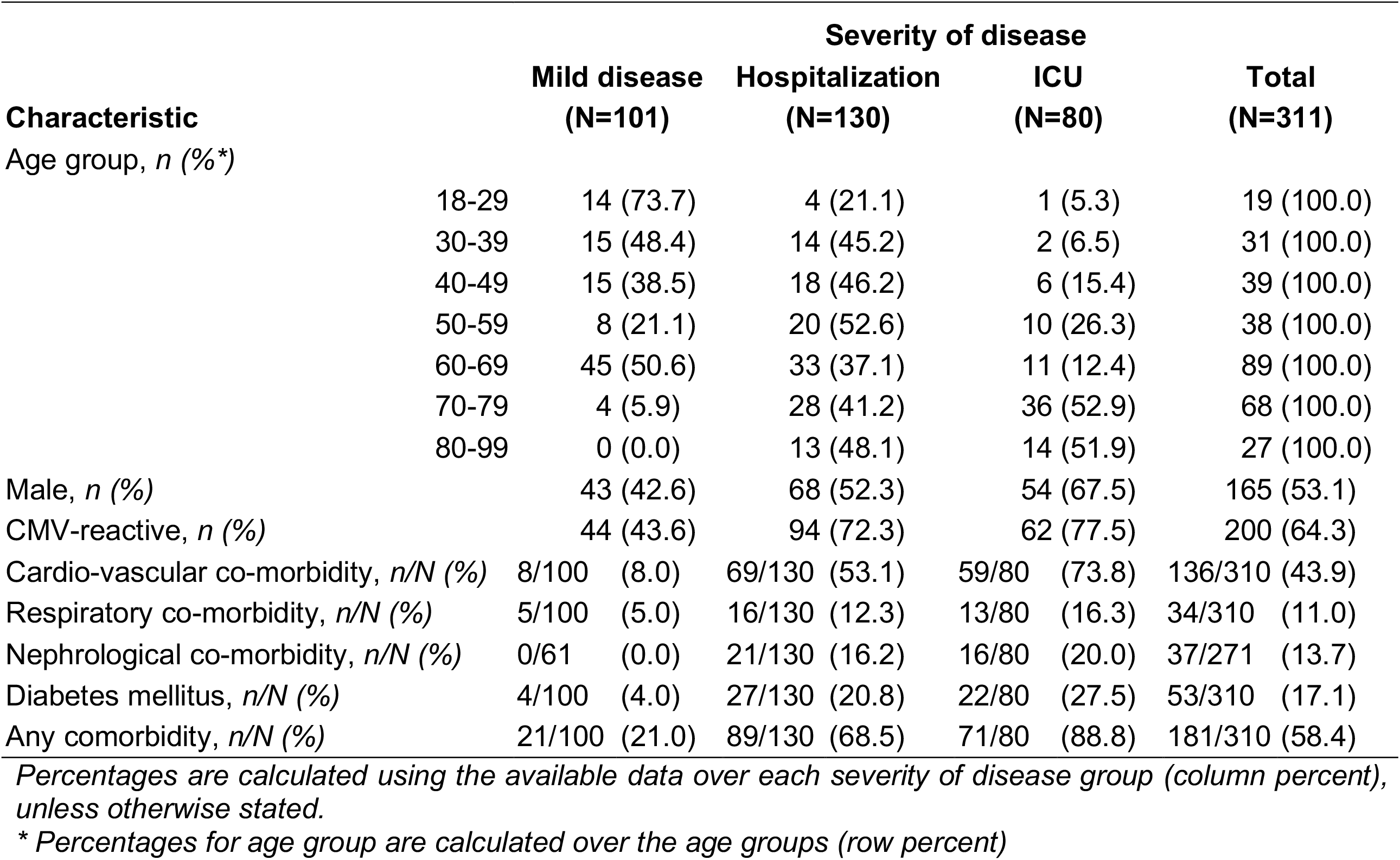
Patient characteristics.

| Characteristic | Severity of disease |  |  |  | Total<br>(N=311) |
| --- | --- | --- | --- | --- | --- |
|  | Mild disease<br>(N=101) | Hospitalization<br>(N=130) | ICU<br>(N=80) |  |  |
| Age group, n (%*) |  |  |  |  |  |
| 18-29 | 14 (73.7) | 4 (21.1) | 1 (5.3) |  | 19 (100.0) |
| 30-39 | 15 (48.4) | 14 (45.2) | 2 (6.5) |  | 31 (100.0) |
| 40-49 | 15 (38.5) | 18 (46.2) | 6 (15.4) |  | 39 (100.0) |
| 50-59 | 8 (21.1) | 20 (52.6) | 10 (26.3) |  | 38 (100.0) |
| 60-69 | 45 (50.6) | 33 (37.1) | 11 (12.4) |  | 89 (100.0) |
| 70-79 | 4 (5.9) | 28 (41.2) | 36 (52.9) |  | 68 (100.0) |
| 80-99 | 0 (0.0) | 13 (48.1) | 14 (51.9) |  | 27 (100.0) |
| Male, n (%) | 43 (42.6) | 68 (52.3) | 54 (67.5) |  | 165 (53.1) |
| CMV-reactive, n (%) | 44 (43.6) | 94 (72.3) | 62 (77.5) |  | 200 (64.3) |
| Cardio-vascular co-morbidity, n/N (%) | 8/100 (8.0) | 69/130 (53.1) | 59/80 (73.8) |  | 136/310 (43.9) |
| Respiratory co-morbidity, n/N (%) | 5/100 (5.0) | 16/130 (12.3) | 13/80 (16.3) |  | 34/310 (11.0) |
| Nephrological co-morbidity, n/N (%) | 0/61 (0.0) | 21/130 (16.2) | 16/80 (20.0) |  | 37/271 (13.7) |
| Diabetes mellitus, n/N (%) | 4/100 (4.0) | 27/130 (20.8) | 22/80 (27.5) |  | 53/310 (17.1) |
| Any comorbidity, n/N (%) | 21/100 (21.0) | 89/130 (68.5) | 71/80 (88.8) |  | 181/310 (58.4) |

*Percentages are calculated using the available data over each severity of disease group (column percent), unless otherwise stated.*
*\* Percentages for age group are calculated over the age groups (row percent)*

**Table 2:** Multinomial logistic regression with dependent variable severity of disease.

| Covariates | Severity of disease |  |  |  |  |  |
| --- | --- | --- | --- | --- | --- | --- |
|  | Hospitalization |  |  | ICU |  |  |
|  | OR | (95% CI) | p | OR | (95% CI) | p |
| **Univariate models:** |  |  |  |  |  |  |
| CMV-reactive | 3.4 | (2.0, 5.9) | < 0.001 | 4.5 | (2.3, 8.6) | < 0.001 |
| Age group | 1.4 | (1.2, 1.6) | < 0.001 | 2.1 | (1.7, 2.7) | < 0.001 |
| Male | 1.5 | (0.9, 2.5) | 0.433 | 2.8 | (1.6, 5.2) | 0.001 |
| Cardio-vascular co-morbidity | 13.0 | (5.8, 29.0) | < 0.001 | 32.3 | (13.4, 77.7) | < 0.001 |
| Respiratory co-morbidity | 2.7 | (1.0, 7.5) | 0.065 | 3.7 | (1.3, 10.8) | 0.018 |
| Nephrological co-morbidity * | 24.2 | (1.3, 433.4) | 0.031 | 31.5 | (1.7, 584.0) | 0.021 |
| Diabetes mellitus | 6.3 | (2.1, 18.6) | 0.001 | 9.1 | (3.0, 27.7) | < 0.001 |
| Any comorbidity | 8.2 | (4.5, 15.0) | < 0.001 | 29.7 | (12.8, 69.0) | < 0.001 |
| **Multivariate model 1:** |  |  |  |  |  |  |
| CMV-reactive | 3.6 | (2.0, 6.4) | < 0.001 | 5.6 | (2.7, 11.5) | < 0.001 |
| Age group | 1.4 | (1.2, 1.7) | < 0.001 | 2.2 | (1.7, 2.9) | < 0.001 |
| **Multivariate model 2:** |  |  |  |  |  |  |
| CMV-reactive | 3.7 | (2.0, 7.0) | < 0.001 | 6.0 | (2.7, 13.5) | < 0.001 |
| Age group | 1.1 | (0.9, 1.4) | 0.278 | 1.6 | (1.2, 2.1) | 0.001 |
| Any comorbidity | 7.4 | (3.7, 14.8) | < 0.001 | 18.1 | (7.1, 46.0) | < 0.001 |

*The reference category is: Mild disease.*
*\* Firth Penalized Likelihood correction in two separate binary logistic regression models due to quasi-complete separation of the data; Firth's correction is not yet implemented for multinomial regression.*

Most interestingly, CMV serostatus was also associated with higher COVID-19 severity, but did not strongly associate with age. CMV-seropositive individuals were more likely to be hospitalized or admitted to ICU (Table 1), and had an increased risk (ORHosp = 3.4, OR_ICU_ = 4.5; both p<0.001) of developing severe COVID-19 (Table 2, univariate analyses). While we observed a tendency towards increasing percentages of CMV-seropositive individuals according to age, we did not find a dominance of CMV-positive over CMV-negative individuals in older (>70 years) compared to younger (<70 years) subjects (Supplementary Fig. 2B). This effect was different from known comorbidities (Supplementary Fig. 2A).

These observations suggested CMV serostatus as a risk factor independent of age. In support of this interpretation, CMV seropositivity remained a significant predictor of unfavorable prognosis after including age (OR_Hosp_ = 3.6, OR_ICU_ = 5.6; both p<0.001) and comorbidities (OR_Hosp._= 3.7, OR_ICU_ = 6.0; both p<0.001) in the multinomial logistic regression model (Table 2, multivariate models).

Looking at CMV serostatus within different disease severities and decades of age further demonstrates that particularly younger patients who required admission to the ICU were mostly CMV seropositive, while this finding weakened with increasing age (Fig. 1). Remarkably, all but one patient younger than 70 years admitted to the ICU and most hospitalized patients were CMV seropositive. Conversely, the CMV prevalence in the mild disease subgroup was similar to the age-matched healthy population in Germany^28^.

**Figure 1.**
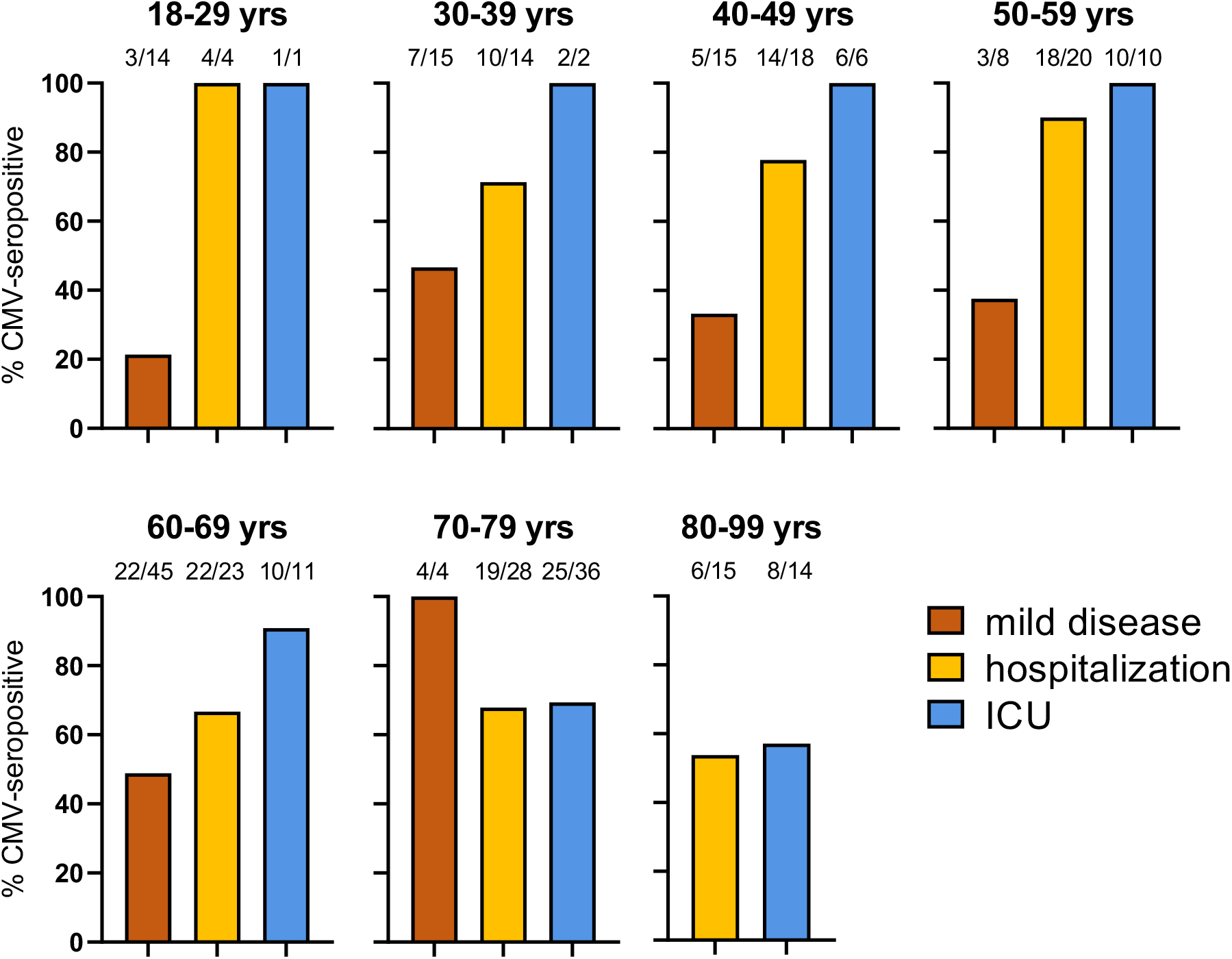
CMV serology associates with severity of COVID-19 in young individuals. CMV IgG titers were measured in serum collected from COVID-patients that either suffered from mild disease or required hospitalization COVID-19 (ICU and non-ICU). Shown are percentages of CMV-positive individuals according to age and disease severity. Numbers above bars indicate the absolute number of CMV-positive subjects on the total number of individual per subgroup.

Classification tree models are known for their ability to identify and graphically display interactions between predictors in a straighter forward way than logistic regression. Important to us was the ability of those models to branch different subpopulations (younger *versus* older patients) using different predictors. Thus, we built the tree-counterpart of the multivariate multinomial logistic model 1 from Table 2 (Fig. 2). Our study cohort was first split according to age and, secondly, only individuals younger than 70 years were further divided according to CMV status. Again, the CMV-positive subgroups (Node 6, 8 and 10) contained a high percentage of patients showing moderate (hospitalized) to critical (ICU) COVID-19 severity (Node 6: 71.1% vs Node 5: 21.6%; Node 8 90.4% vs Node 7 28.6%; Node 10 59.2% vs Node 9 34.3%). Furthermore, CMV serostatus reached increasing odds ratio with decreasing age (OR (95%CI) for age <=49: OR_Hosp_ = 5.6 (2.4, 17.0), OR_ICU_ = 16.3 (2.0, 663.5); age 50-59: OR_Hosp_ = 7.5 (1.8, 76.3), OR_ICU_ = 12.5 (1.4, 760.6); age 60-69: OR_Hosp_ = 1.8 (0.8, 5.1), OR_ICU_ = 5.0 (1.2, 44.5)), mainly due to a more pronounced difference in the hospitalization rate between CMV-positive and CMV-negative individuals (Fig. 2).

**Figure 2.**
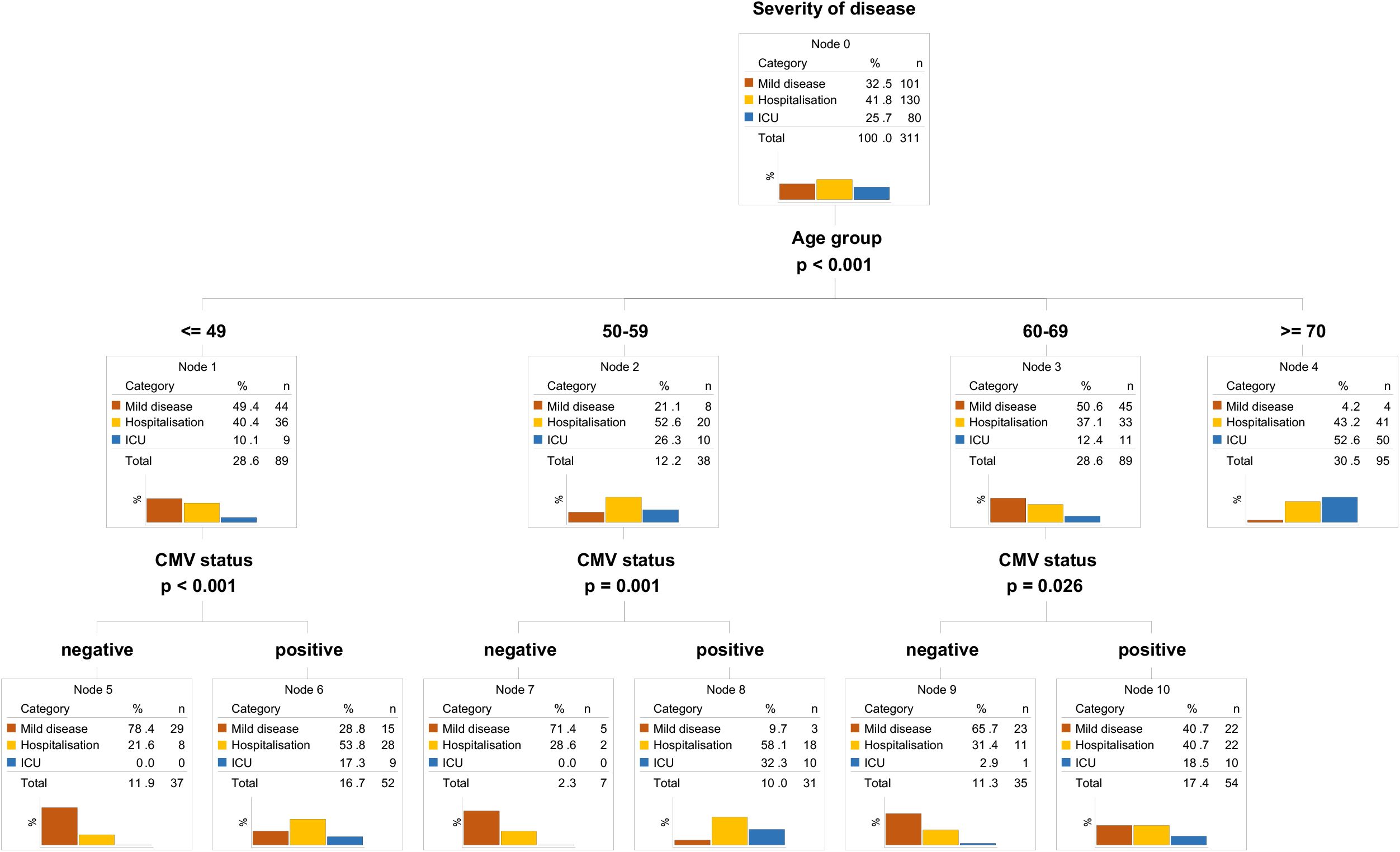
CMV serostatus predicts outcome in young individuals. Classification tree model (CHAID) using CMV serostatus and age as predictors of severity of disease.

In a second classification tree model we further analyzed the predictive value of CMV serostatus in relation not only to age but also to the available comorbidities. As expected, having a known comorbidity was a predominant indicator of poorer prognosis especially for the very old, as most of the ICU patients were found in this group (Fig. 3, Node 1). Intriguingly, after age stratification, younger patients suffering from comorbidities (<70, Fig 3, node 3) were more likely to develop a severe course of disease requiring ICU treatment when CMV-seropositive (CMV positive: 33.3%; CMV negative: 4.0%) (Fig 3, nodes 7 and 8). In individuals without known co-morbidities, CMV seropositivity again served as a negative predictor of outcome, but was independent of age (node 5 and 6).

**Figure 3.**
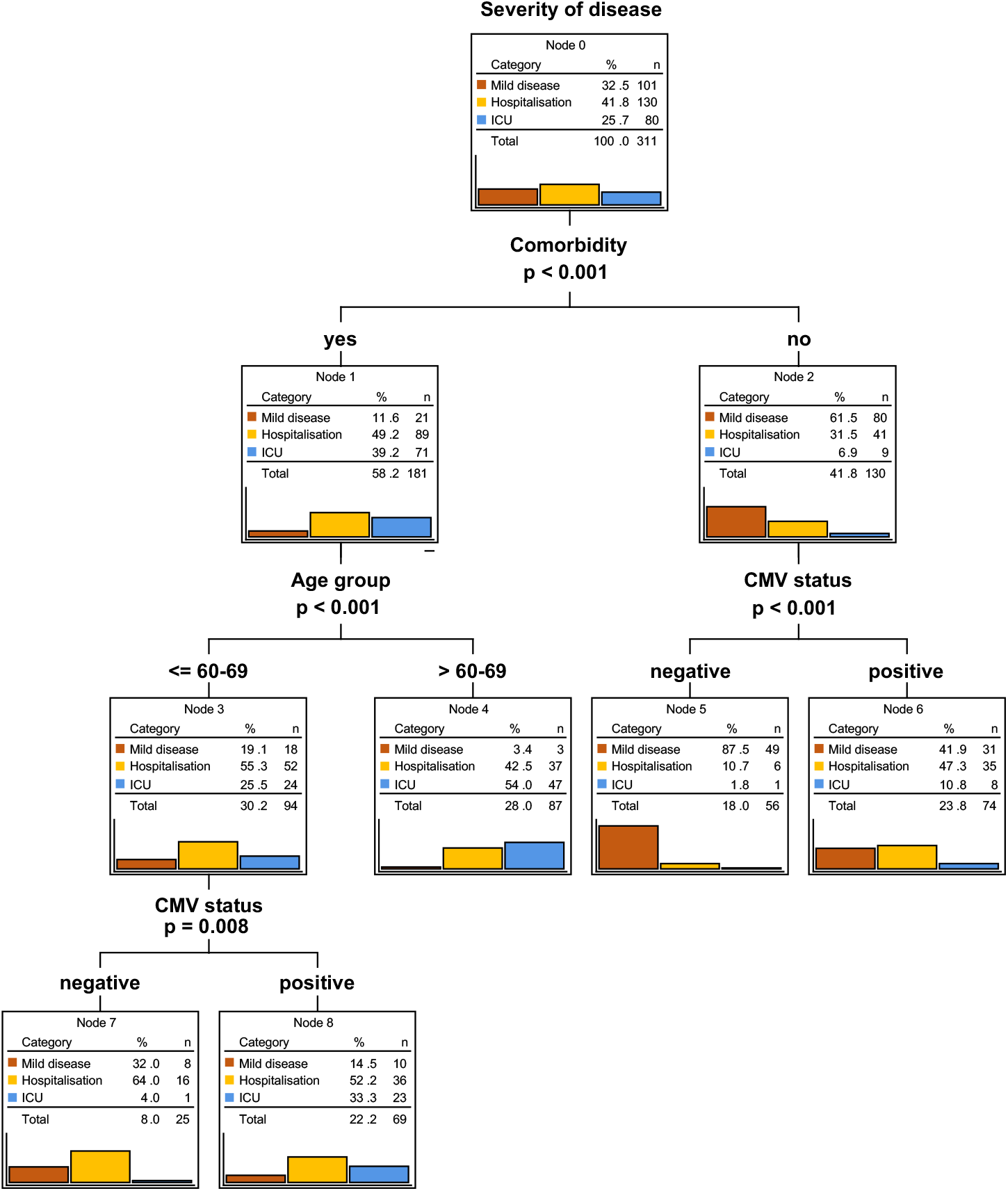
CMV serostatus remains an independent predictor of worse outcome in presence of comorbidity. Classification tree model (CHAID) using CMV serostatus, comorbidities and age as predictors. Information on comorbidities is missing for one individual that was included into the no comorbidities group for practicability.

Overall, our data raise evidence that CMV serostatus might be a very strong and independent risk factor for severe COVID-19, particularly in younger individuals.

## DISCUSSION

In this study, we identified ‘CMV-seropositivity’ as a potential novel risk factor for severe COVID-19 in individuals younger than 70 years. Our current data cannot distinguish whether CMV-seropositivity is just a biomarker or more directly involved in the pathophysiology of severe COVID-19 in younger individuals. Further research in this direction should be rapidly performed, as the underlying mechanisms might also open up novel options for therapy improvement.

The identification of CMV/SARS-CoV-2 cross-reactive T cells (Suppl. Fig. 1) might indicate that CMV infection is indirectly involved in severe COVID-19 via the preferential recruitment of T cells from the antigen-experienced or memory T cell pool. Such T cells are often less reactive to the antigen for which they were not originally primed and because of this an impaired T cell response could fail to control SARS-CoV-2, thereby leading to severe COVID-19. Due to the phenomenon of ‘memory inflation’, CMV-specific T cells often dominate the general memory T cell population, especially in older CMV-seropositive individuals where the pool of naïve T cells narrows. Therefore, CMV-specific T cells might have a higher likelihood of participating in the pool of recruited SARS-CoV-2 specific T cells from cross-reactive repertoires. But this phenomenon is certainly not restricted to CMV. Cross-reactivity to SARS-CoV-2 epitopes in severe COVID-19 patients has also been shown for other target specificities, such as other common cold corona viruses^29–35^. Many groups worldwide, including ourselves, are currently trying to shed more light on the relevance of recruitment of SARS-CoV-2-specific T cells from cross-reactive antigen-experienced T cell repertoires for severe COVID-19, and CMV might be a “master factor” in this context considering its extreme impact on T cell repertoire shifts. However, with the existing body of data demonstrating that CMV supports immunosenecence especially in elderly individuals, it remains surprising that our current study on COVID-19 identified a correlation between CMV seropositivity and disease severity particularly for younger patients. If CMV seropositivity would indeed impair the quality of SARS-CoV-2 specific T cells responses in severe COVID-19, adoptive T cell therapy with highly SARS-CoV-2-specific T cells might become an interesting option to therapeutically compensate for the defect. Indeed, first clinical trials in this direction are currently ongoing and recent trials based on adoptive transfer of memory T cells from convalescent donors have shown some promising results^36^.

A completely different scenario would be a more direct involvement of CMV in severe COVID-19 pathogenesis of in younger individuals via CMV reactivation. Few recent case reports have described CMV-reactivation during SARS-CoV-2 and postulated that CMV-driven pneumonitis might have been a key driver of lung function compromise and clinical outcomes in these COVID-19 patients^16,17,19^. Pathophysiologically, inflammatory cytokines stimulated by SARS-CoV-2 could lead to the reactivation of latent CMV residing in the lung. We have tried searching retrospectively in our cohort for evidence of CMV reactivation (e.g. via CMV PCR in bronchoalvelolar lavages; data not shown), but so far failed to demonstrate more clear evidence for reactivation. Unfortunately, these results are not conclusive, since demonstration of CMV reactivation is complex and requires optimal sample acquisition and diagnostics. We are currently initiating prospective studies to specifically search for evidence of CMV reactivation during severe COVID-19.

Although our study shows surprising results that are possibly impactful for COVID-19 patients’ outcomes, there are also some limitations that should be mentioned. Our cohort comprises patients and biological samples that were collected in Germany earlier in the pandemic. Therefore, it is important to initiate similar studies with additional subjects to confirm whether our findings can be generalized to patients from other countries. Also, socioeconomical factors should be taken into consideration. Additionally, the biomaterial was collected before the emergence of variants of concern that are currently dominating the pandemic (e.g. delta variant in Europe) and before the global vaccination campaign. Thus, it will be important to perform follow-up analyses in settings that also render the current infection and vaccination dynamics. Another limitation of our study is that the different patient subgroups are not fully balanced by age and gender – which is partly due to biological reasons (for example absence of mildly symptomatic elderly individuals > 80 years). As the biomaterial and patient data used for our analyses were collected in the context of different study protocols, availability of data varied. All of these factors added some challenges to the statistical analyses; however, despite these limitations, the main findings summarized in this report remain robust and highly significant.

In summary, we identified ‘CMV-seropositivity’ as a novel risk factor for severe COVID-19 in younger individuals. Our findings may have immediate implications on patient management and inspire investigation into SARS-CoV-2 vaccine response quality with respect to CMV serostatus in more detail.

## Data Availability

All data produced in the present study are available upon reasonable request to the authors

## AUTHOR CONTRIBUTIONS

E.D., D.H.B., S.W. conceptualized the study; S.W. performed experiments and data analyses; V.K. performed and described statistical analyses; T.B., P.S., M.A., C.S.C. collected samples and clinical information of mild COVID-19; J.E., C.W, S.D.S designed and organized the study on hospitalized COVID-19 patients; A.M.J., D.H. and U.P. performed CMV serology measurements; K.I.W., K.S., A.M.J. provided resources; M.G, D.H.B, E.D. acquired funding; E.D., V.K. prepared figures and tables; S.W., D.H.B wrote the manuscript; D.H.B. and E.D. supervised the study and administered the project. All authors read and approved the manuscript.

## ACKNOWLEDGEMENTS

This study was supported by the EIT Health CoViproteHCt #20877 and the German National Network of University Medicine of the Federal Ministry of Education and Research (BMBF; NaFoUniMedCovid19, 01KX2021; COVIM).

## MATERIAL AND METHODS

### Clinical samples

For mildly symptomatic SARS-CoV-2 infections, blood samples were collected at the Helios Klinikum München West (n = 39), from healthcare employees who were diagnosed via PCR and experienced mild symptoms (cold, cough and mild fever), but did not require hospitalized treatment at any time. Additional biosamples from mildly diseased patients were acquired from the university hospital Köln in the context of the Nationales Netzwerk Universitätsmedizin consortium (n = 62). Hospitalized patients (ICU, n= 80 and non-ICU, n=130) were prospectively included in the COVID-19 registry COMRI at the University Hospital rechts der Isar. Serum samples were collected according to the study protocol. Clinical data were retrospectively collected by medical chart review.

All participants provided informed written consent. Approval for the study design and sample collection was obtained from the local ethics committee of the Technical University of Munich (reference number 182/20 and 633/21 S-SR) and the COVIM steering committee.

### Cell isolation and culture conditions

PBMCs were isolated from whole blood by gradient density centrifugation according to manufacturer’s instructions (Pancoll human) and either frozen at -80 °C in a freezing medium composed of 90% FCS and 10% DMSO. PBMCs were cultured in RPMI 1640 supplemented with 10% FCS, 0.025% l-glutamine, 0.1% HEPES, 0.001% gentamycin, 0.002% streptomycin and 180 U/ml IL-2 in a humidified incubator at 37 °C and 5% CO_2_.

### TCR DNA template design and CRISPR/Cas9-mediated TCR knock-in

DNA constructs for CRISPR/Cas-9-mediated HDR at TRAC locus were designed *in silico* with the following structure: 5′ homology arm (300–400 base pairs), P2A, TCR-β (including mTRBC with additional cysteine bridge), T2A, TCR-α (including mTRAC with additional cysteine bridge), bGHpA tail, 3′ homology arm (300–400 base pair). All HDR DNA template sequences were synthesized by Twist.

CRISPR/Cas9-mediated endogenous TCR knock-out and transgenic TCR knock-in (KI) was performed as described ^5^. Briefly, freshly isolated PBMCs were activated with CD3/CD28 Expamer (Juno Therapeutics), 300 U/ml IL-2, 5 ng/ml IL-7 and 5 ng/ml IL-15. After removing of the stimulus by incubation in a Biotin solution, cells were electroporated in a Nucleofector Solution containing Cas9 ribonucleoprotein and DNA templates with a 4D Nucleofector XL unit (Lonza). After electroporation, cells were cultured in RPMI with 180 IU/ml IL-2 before analysis.

### Intracellular cytokine staining

TCR-engineered PBMCs were stimulated with the peptide pool of interest (PepTivator® SARS-CoV-2 Prot_S from Miltenyi Biotech or PepMix™ HCMVA (pp65) from JPT) at a concentration of 1 µg/ml. For TCR-engineered T cells, autologous antigen presenting cells (PBMCs) were loaded with the different peptide mixes via incubation for 2 h at 37 °C, and co-cultured with engineered T cells in a 1:1 effector:target ratio. Unpulsed PBMCs served as negative control whereas 25 ng/ml PMA and 1 µg/ml Ionomycin served as positive control. After incubation for 4 h at 37 °C in presence of 1 µg/ml GolgiPlug (Brefeldin A), cells were stained with EMA solution (1:1000) for live/dead discrimination and subsequently with surface antibodies: CD8-PE (1:200), CD3-BV421 (1:100) and murine TCR β-chain-APC/Fire750 (1:50). Cells were fixed using Cytofix/Cytoperm solution followed by staining for intracellular cytokines by IFN-γ-FITC antibody (1:10) and IL-2-APC (1:25). Flow cytometric analysis was performed on the CytoFlex S Cell Analyzer.

### CMV serology

Analyses were conducted at the Institute for Virology, Technical University Munich. CMV IgG was measured in serum samples with a chemiluminescent microparticle immunoassay on Architect i1000 (Abbott GmbH, Wiesbaden). The cut-off value was 6 AU/ml.

### Statistical methods

Descriptive statistics are provided as absolute and relative frequencies by severity of disease and in total. Information about patient age was collected on an ordinal scale. Univariate and multivariate multinomial logistic regression models were calculated using “mild disease” as reference category of the dependent variable severity. Due to quasi-complete separation of the data, some models needed a Firth Penalized Likelihood correction. This solution is available only for the binary logistic regression, which is why the two binary logistic regressions were calculated instead of one multinomial logistic regression. The odds ratios (OR) are presented together with their 95% CI and the corresponding p-value. In addition, classification tree models (CHAID) were built from all available data using the following specifications: dependent variable severity of disease, pearson chi^2^ statistic for the split, Bonferroni-adjusted p-values, 10-fold cross validation, and minimum number of cases in a parent node 20; in a child node 7. The significance level was set to 5%. Analysis was performed using IBM SPSS version 26 (IBM Corp., Armonk, N.Y., USA) and SAS 9.4 (SAS Institute Inc., Cary, NC, USA).

## FIGURE LEGENDS

**Supplementary Fig. 1.**
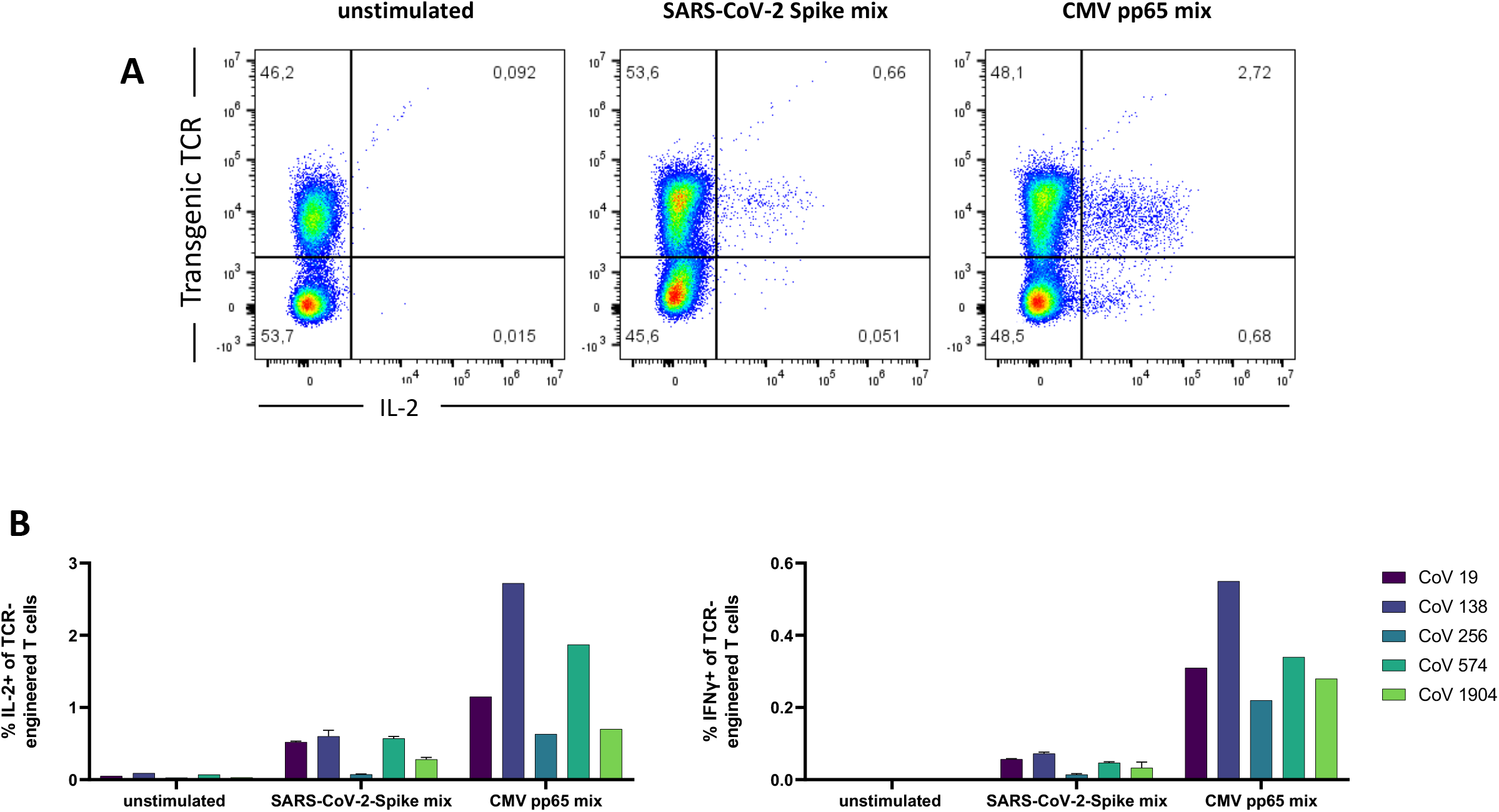
Cross-reactivity of SARS-CoV-2-specific TCRs to CMV. TCRs were isolated from an ICU patient and engineered into PBMCs from healthy donors via CRISPR/Cas9-mediated knock-in. Engineered T cells were co-cultured with autologous PBMCs previously pulsed with 1 µg/ml Peptivator S mix or CMV pp65 mix for 4 h at 37 °C. Shown are representative raw data (A) and quantification (B) of IL-2 and IFN-γ production.

**Supplementary Fig. 2.**
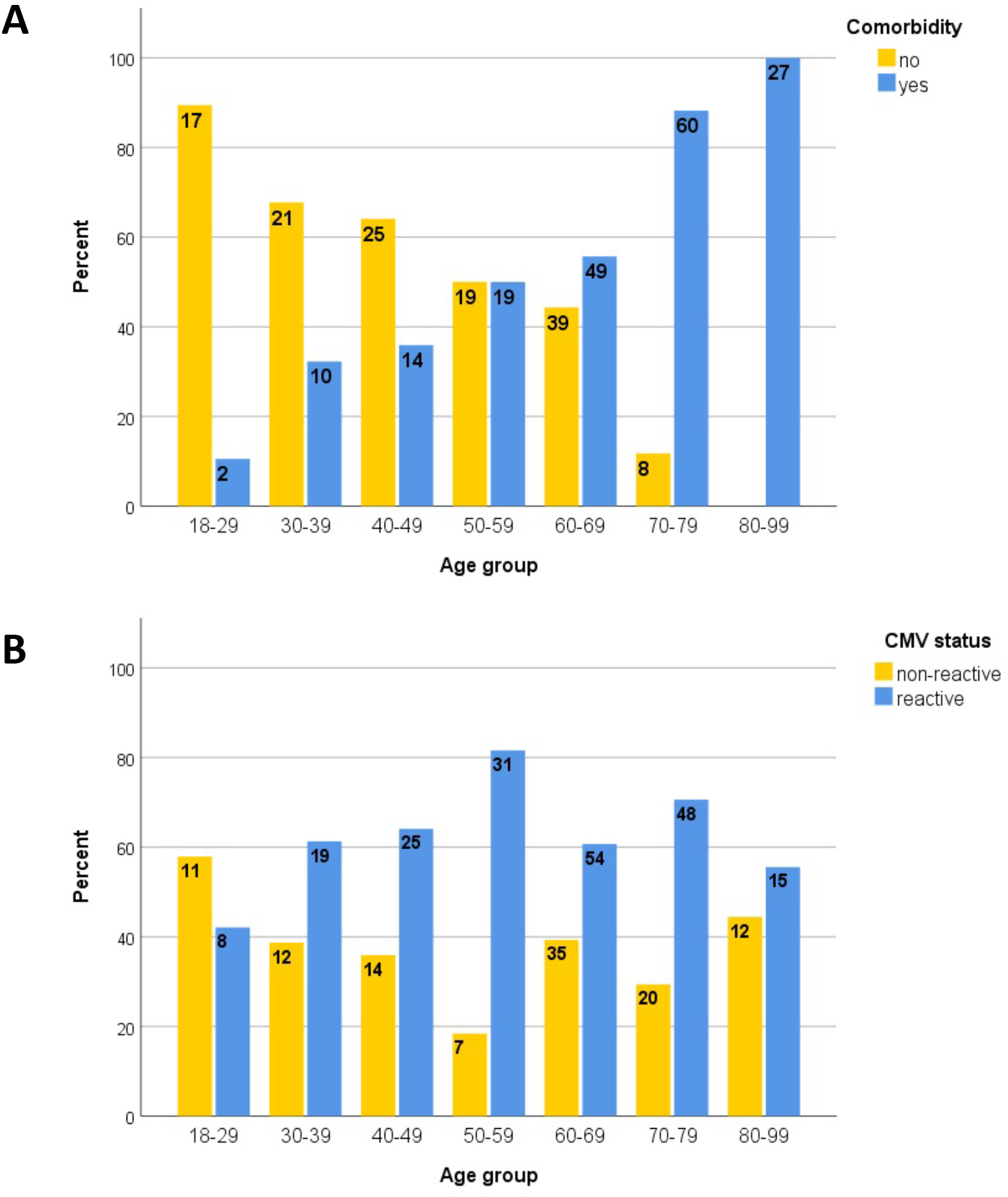
Occurrence of comorbidity and CMV according to age. Bar graphs showing the percentage of individuals enrolled in this study with or without comorbidities (A) and positive or negative CMV serostatus(B). Numbers within the bars indicate absolute numbers of individuals.

